# Human and Viral Whole-Genome Sequencing Reveals HPV-Driven Carcinogenesis and Mutational Signatures in Sinonasal Squamous Cell Carcinoma

**DOI:** 10.64898/2026.02.04.26345593

**Authors:** Lukasz Zareba, Samuli Eldfors, Maoxuan Lin, Michael E. Bryan, William C. Faquin, Lisa Mirabello, Sambit K. Mishra, James S. Lewis, Michael S. Lawrence, Daniel Faden

## Abstract

Sinonasal squamous cell carcinoma (SNSCC) is an aggressive malignancy that has not seen therapeutic advance in decades. Historically attributed to inhaled carcinogens, SNSCC is also incidentally associated with human papillomavirus (HPV), the major oncogenic driver in over 80% of adjacent oropharyngeal cancers. While HPV status guides staging and treatment in oropharyngeal cancer, the oncogenic consequences of host–virus interactions in SNSCC remain incompletely defined. Here, through paired host and viral whole-genome sequencing of 21 cases, we map the genomic footprint of HPV in SNSCC. Strikingly, less-studied genotypes such as HPV45 and 51 constitute driver infections. APOBEC activity was detected in a subset of HPV-associated SNSCC, while Signature 12 emerged as the most frequent dominant mutational signature and showed transcriptional strand bias consistent with an in vivo mutational process. Viral integration events, including extrachromosomal DNA (ecDNA)-associated HPV–human ecDNA amplicons, further contribute to tumor evolution.

**Statement of Significance:** Paired host and viral whole-genome sequencing identifies non-HPV16 high-risk HPV genotypes (e.g., HPV45 and 51) and HPV–human ecDNA amplicons as primary oncogenic drivers of sinonasal squamous cell carcinoma (SNSCC). A distinct mutational landscape, characterized by frequent Signature 12 activity and APOBEC mutagenesis in a subset of tumors, distinguishes SNSCC from other HPV-associated cancers. Importantly, non-HPV16 high-risk HPV genotypes play an underrecognized role in SNSCC, and support their inclusion in routine clinical testing.

## Introduction

Sinonasal squamous cell carcinoma (SNSCC) is an aggressive head and neck squamous cell carcinoma (HNSCC) of the sinonasal cavity that has not seen major therapeutic advances in decades. Though historically attributed to inhaled carcinogens such as hardwood dust and tobacco smoking, SNSCC is incidentally associated with human papillomavirus (HPV). Importantly, HPV is the primary oncogenic driver of >80% of anatomically adjacent oropharyngeal cancers(^1^). Our group and others have demonstrated that HPV is detectable in nasopharyngeal and sinonasal cancers(^2^), possibly due to refluxed secretions from the oropharynx(^3^). Notably, these cancers are rising both in incidence and within the same geographic locale as HPV+OPC, the most common HPV-associated cancer in the US(^3^). Previous profiling suggests that HPV-associate sinonasal squamous cell carcinoma (HPV+SNSCC) shares mutational profiles with HPV-associated cervical and HPV+OPC(^4^). However, definitive genomic mapping of potentially oncogenic host-virus interactions through systematic sequencing of paired tumor-normal specimens remains un-explored. Here, we leverage joint whole-genome sequencing (WGS) of both human and HPV genomes to interrogate the mutational signatures and structural events underpinning viral oncogenesis in SNSCC.

## Results

Using a cohort of 21 SNSCC paired tumor-normal specimens, we subjected each sample to orthogonal tumor WGS (mean depth, 59x), matched normal WGS (mean depth, 30x), ultra-deep HPV genome WGS (10,000x), and a broad HPV DNA PCR panel (see Methods). Per-sample, clinicopathologic, sequencing, and HPV assay results are provided in Supplementary Table S1. Eighteen of 21 (86%) cases were HPV-associated by human WGS and/or HPV DNA PCR, with 94.4% concordance between the two technologies (17/18 jointly profiled; 95% CI, 74.2–99.0%; binomial *P* < 0.001; see Methods). Genotype concordance was 81.0% (17/21; 95% CI, 60.0–92.3%; binomial *P* = 0.001). Sixteen of 21 (76%) were p16-positive, a surrogate marker for HPV-infection in the oropharynx (Table 1). Among patients with a smoking history, 7/14 (50%) had ≥10 pack years (Table 1). Unlike HPV+OPC, where HPV16 predominates (∼90%)(^5^) only 6/18 (33%) HPV+SNSCC had single-genotype HPV16 (Fig. 1a). Non-HPV16 high-risk genotypes were present in 10/18 (55%) HPV+SNSCC patients, a rate substantially higher compared to HPV+OPC (∼12.5%)(^6^) (Fig. 1a). We observed eight distinct HPV genotypes (Fig. 1a), including one multi-genotype infection and two tumors with low-risk HPV11 infection. Both low-risk (e.g. HPV11) infections and a plurality of high-risk genotypes which vary in prevalence across human cancers (e.g. HPV 16, 18, 35, 39, 45, 51, 59) were present. Sequencing coverage metrics for HPV genomes, including depth and breadth across detected genotypes, are summarized in Supplementary Table S2. Interestingly, a broad representation of high-risk HPV genotypes other than HPV16 has also been observed in nasopharyngeal cancer, an oropharynx-adjacent site where HPV also appears to play a role(^7^).

**Figure 1.**
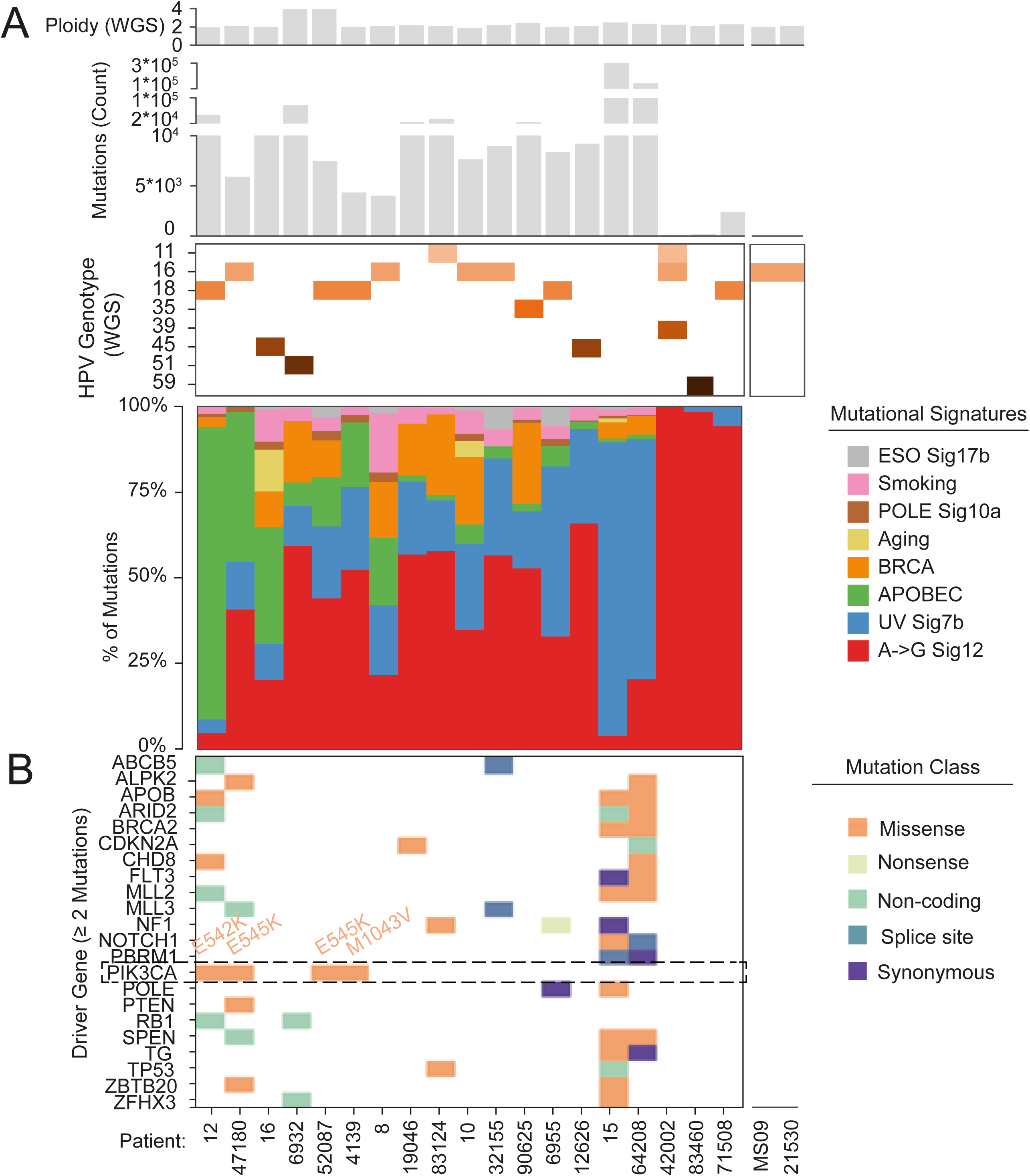
A portrait of genomic aberrations in SNSCC. (a) Oncoplot summarizing the genomic landscape of 21 SNSCCs, including: tumor ploidy, overall mutational burden, HPV genotype by WGS, and the relative contribution of different single base substitution signatures. (b) Point mutations in known cancer driver genes, highlighting APOBEC-induced *PIK3CA* driver mutations.

**Table 1.** Patient Demographics and Clinical Characteristics of the.

| Characteristic | N = 21 |
| --- | --- |
| **Demographics** |  |
| Age at diagnosis, years — median (range) | 72 (54–96)† |
| **Sex** |  |
| Male | 13 (81) |
| Female | 3 (19) |
| Unknown/Missing | 5 |
| **Tumor Characteristics** |  |
| **Tumor site** |  |
| Nasal Cavity | 12 (75) |
| Maxillary Sinus | 3 (19) |
| Nasal Cavity OR Maxillary Sinus | 1 (6) |
| Unknown/Missing | 5 |
| **Histopathology** |  |
| Non-Keratinizing | 18 (86) |
| Keratinizing | 3 (14) |
| **Inverted papilloma association** |  |
| Yes | 9 (56) |
| No | 7 (44) |
| Unknown/Missing | 5 |
| **HPV and Biomarker Status** |  |
| **HPV status (WGS)** |  |
| Positive | 18 (86) |
| Negative | 3 (14) |
| **HPV genotype (among HPV-positive)** |  |
| HPV16 | 8 (44) |
| HPV18 | 5 (28) |
| HPV35 | 1 (6) |
| HPV39 | 1 (6) |
| HPV45 | 1 (6) |
| HPV51 | 1 (6) |
| HPV59 | 1 (6) |
| **p16 IHC** |  |
| Positive | 16 (76) |
| Negative | 5 (24) |
| **Smoking history** |  |
| ≥10 pack-years | 7 (33) |
| <10 pack-years | 7 (33) |
| Unknown/Missing | 7 (33) |

patients.
Percentages calculated among patients with available data for each variable.
HPV status determined by whole-genome sequencing (WGS) of tumor samples.
HPV genotype reflects the dominant type detected by WGS.

Integration of all or part of the HPV genome is a well-described carcinogenic mechanism in HPV+OPC, as well as cervical cancer. We identified high-confidence integration breakpoints in most HPV+ tumors, with as many as 7 distinct breakpoints per tumor. In total, 28 unique, high-evidence viral-host junctions were mapped (Fig. 2a, Supplementary Tables S1–S5). In the one case harboring multi-genotype infections, only a single dominant integrating genotype was observed, suggesting that one genotype primarily drives oncogenic progression, even amid co-infection by other HPV types. This observation is consistent with findings in cervical cancer(^8^). One HPV11+ tumor lacked any high-risk genotype and showed no evidence of viral integration but harbored a *TP53* mutation and was p16-negative, consistent with HPV11 as a bystander in this case (Supplementary Tables S6-S8).

**Figure 2.**
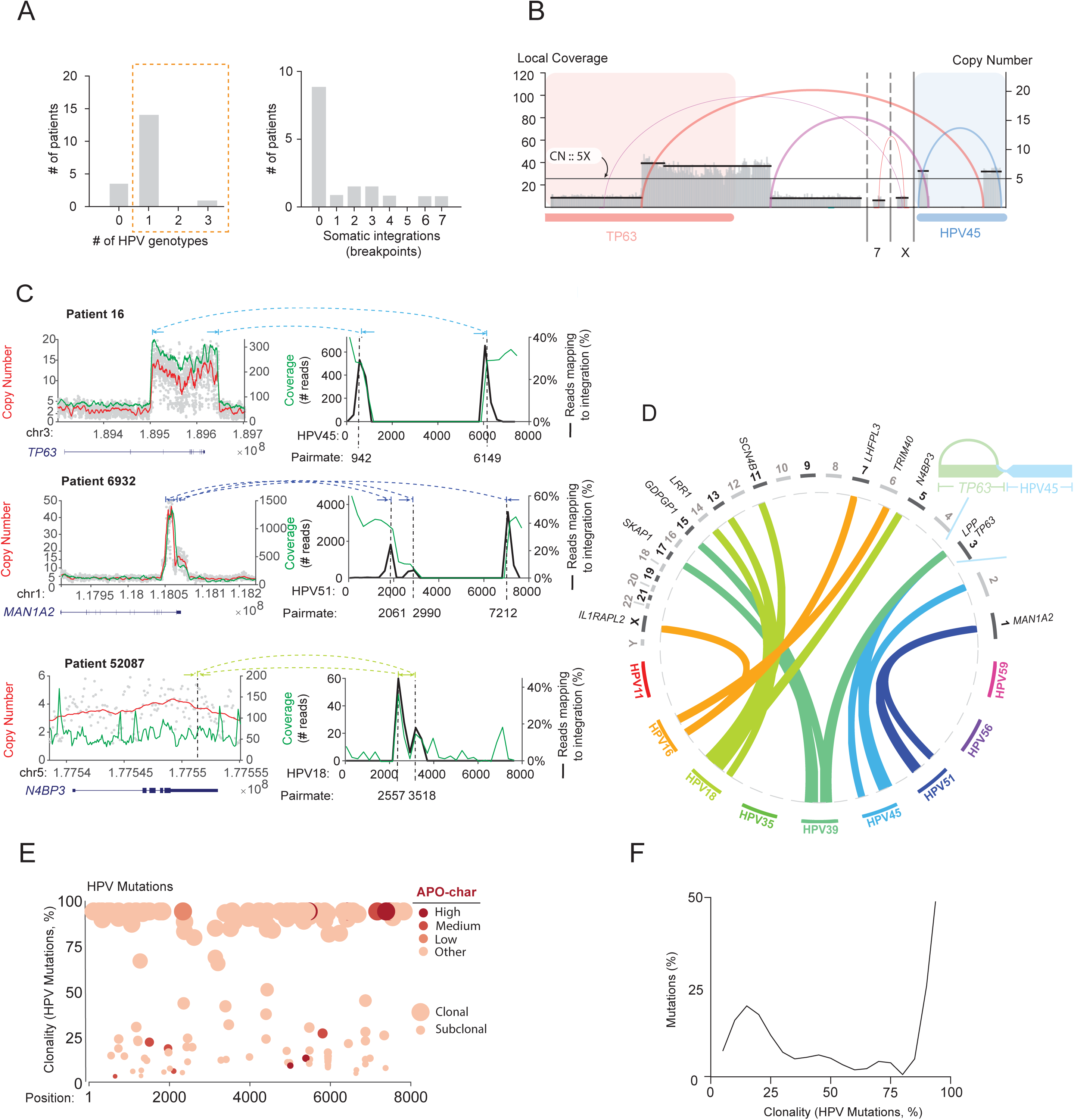
Genomic record of HPV activity in SNSCC. (a) Distribution of HPV positivity (total number of genotypes per patient) and detectable hg19-HPV breakpoints among HPV+SNSCC patients. (b) Focal segmentation of HPV-encoding extrachromosomal DNA hijacking the *TP63* oncogene, resulting in local copy number gain. A semi-circular arc connecting two loci denotes chimeric reads which map to two sites. (c) High-resolution characterization of select HPV integration sites in 3 HPV+SNSCC patients, showing the human locus (left) and the corresponding HPV genome (right). Copy number segmentation (red) is overlaid on per-bin copy number estimates (grey) against hg19 coordinates, with local read depth in green. Within the HPV genome, viral coverage (green) is plotted against the proportion of reads spanning a viral-host junction (black). Pairmate values give the HPV coordinates of each breakpoint. A dashed arc connecting two loci denotes the human and viral breakpoints of a single chimeric junction. Co-option of the *TP63* oncogene into an HPV45-containing amplicon (top). Focal copy number gain (50X genomic median) of *MAN1A2*-HPV51 amplicon (middle). Evidence of subclonal integration of HPV18 (3′ UTR of *N4BP3*) without corresponding copy number instability in the human genome (bottom). (d) Circos plot mapping the somatic integration of HPV into the human genome, sub-stratified by HPV genotype and highlighting the identification of a chimeric *TP63*-HPV45 amplicon. Each arc represents chimeric reads mapping across two sites. (e) Clonality of mutations across the HPV genome as detected by viral WGS, highlighting the contribution of APOBEC mutagenesis. (f) Distribution of mutational clonality within the HPV genome.

Our cohort also exhibited novel patterns of HPV integration (Fig. 2b,c,d). In several cases, integration was accompanied by profound local copy-number amplification of host and viral sequences. Chimeric reads revealed an extrachromosomal DNA (ecDNA) species fusing viral DNA from HPV45, a rare genotype in HPV+OPC, to host DNA from the 3′ region of the *TP63* oncogene (Fig. 2b, Supplementary Table S3). Focal segmentation of the somatic breakpoints delimiting this fusion revealed a ∼50kbp segment of chromosome 3q which achieved amplification to ∼15–20X genomic median (i.e. “copies”). This segment included the HPV45 *E6* URR sequences fused to *TP63*. The structure and amplitude of this amplicon resemble oncoviral integration-linked ecDNA(^9^) (Fig. 2c, top). To our knowledge, this is the first demonstration of an HPV ecDNA-like amplification in SNSCC.

We further observed amplification in a tumor with HPV51 with three distinct HPV51 breakpoints clustered at the 3′ end of *MAN1A2*. Each breakpoint was associated with a distinct copy-number profile, but all showed high-level amplification (∼15–50 copies). This suggests either sequential chromosomal rearrangements or the presence of multiple “nested” ecDNA species containing segments of HPV51 and *MAN1A2* (Fig. 2c, middle). Initial integration may have been followed by stepwise extrusion of some segments via iterative breakage-fusion cycles. These findings strongly suggest an oncogenic role of less-common HPV genotypes in SNSCC(^10^). Yet not all HPV integrations cause detectable copy-number changes. For instance, one tumor showed evidence of HPV18 integration at the 3′ end of *N4BP3* without an associated copy-number change (Fig. 2c, bottom).

We next interrogated the host somatic mutational landscape. Clonal mutations were observed in common driver genes regulating mitogenic signaling and genomic integrity (e.g. *PIK3CA, KDM6A, MLL3, BCOR, CHEK2*) (Fig. 1b; Supplementary Tables S6–S8). Among high-risk HPV+SNSCC cases, 4/17 (23%) harbored *PIK3CA* missense mutations (Fig. 1b). Three of these were in the helical domain, a hotspot enriched for APOBEC3-mediated mutations(^11^). *TP53* and *CDKN2A* mutations were mutually exclusive with high-risk HPV infection, whereas *PIK3CA* mutations were seen only in high-risk HPV+ cases. These observations are consistent with reports of depletion of *TP53/CDKN2A* mutation in HPV-associated cancers, as well as enrichment of mutations in the PIK3CA helical domain(^12^). FGFR3 S249C mutations, recently noted in SNSCC(^4^), were not detected in this cohort. Genome-wide, SNSCCs exhibited a mutational rate like that of previously profiled HNSCC (Fig. 3a). Despite adequate sequencing depth and sufficient tumor purity, we did not recover any confidently called somatic mutations in two HPV16-positive tumors after stringent filtering (tumor purity MS09 = 64%, 21530 = 46%).

**Figure 3.**
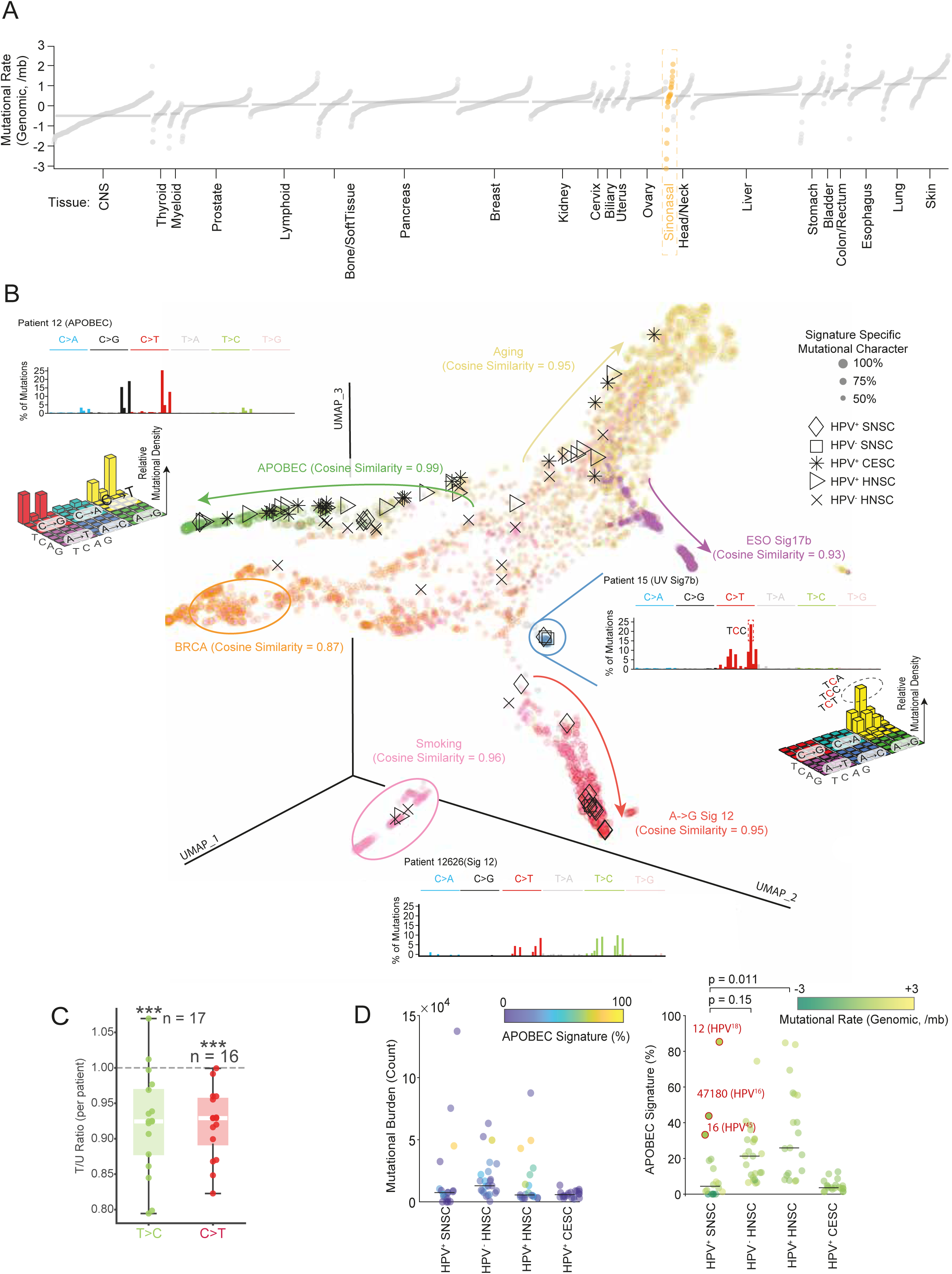
Pan-cancer context of mutational signature activity in SNSCC. (a) Scatterplot depicting the genomic mutational rates of different TCGA-ICGC cancer lineages, highlighting SNSCC. (b) Cartography of mutational signatures across human cancers, highlighting the distribution of SNSCC relative to TCGA HNSCC. UMAP loadings of seven prominent SBS signatures (BRCA, APOBEC, Aging, UV Sig7b, A◊G Sig 12, Smoking) are used to partition individual TCGA-ICGC tumors along different mutational “trajectories.” Each dot in the cartography represents a single tumor and is colored by the dominant mutational signature. Select cancer subtypes (e.g. HPV+HNSCC) are highlighted with symbols. Select examples of patients which are enriched for different SBS signatures e.g. Patient 12626 Sig 12, Patient 12 APOBEC, Patient 15 UV Sig 7b. (c) Transcriptional strand bias in the two major Signature 12 substitution channels. Per-patient T/U ratios are shown for T>C (green) and C>T (red) mutations in SNSCC tumors, representing 65% and 12% of the Sig. 12 spectrum, respectively. Each dot denotes one patient. Box plots indicate the median (white line), interquartile range (box), and whiskers extending to 1.5 × IQR. The dashed line marks T/U = 1.0, the expected ratio for FFPE-derived artifact. P-values by two-sided Wilcoxon signed-rank tests against T/U = 1.0; \*\*\**P* < 0.001. (d) Left, scatter plot directly compares the total mutational burden in HPV+SNSCC to HPV-HNSCC, HPV+HNSCC, and HPV+CESC. Right, scatter plot directly compares APOBEC signature loadings among different HNSCC subtypes, highlighting select SNSCC which are both HPV+ and APOBEC-enriched. P-values were computed using the MATLAB ttest2() function.

Given that APOBEC mutagenesis is a major mutational process in HPV-associated carcinomas(^13^), we interrogated mutational signatures in both host and viral genomes. Mutational signature analysis identified APOBEC mutagenesis in a subset of SNSCC tumors (Fig. 1a). Unexpectedly, Signature 12 (Sig 12) was the most frequent dominant mutational signature in HPV+SNSCC, despite not having been previously reported in this disease (Fig. 1a). Across a pan-cancer cohort of 3,025 tumors spanning 35 tumor types, SNSCC exhibited the highest mean Sig 12 fractional exposure (50.1%), with 90.5% of SNSCC tumors harboring >5% Sig 12 activity (Fig. 3b). Because Sig 12 has no established etiology, and because FFPE processing can distort mutational spectra, we next assessed transcriptional strand bias to test whether the observed pattern was consistent with an *in vivo* mutational process. Evaluable tumors were required to have more than 100 genic mutations assigned to each transcriptional strand for the corresponding strand-bias analysis. Sig 12 substitution channels exhibited significant transcriptional strand bias across the SNSCC cohort (Fig. 3c). In the T>C channel, 15 of 17 evaluable tumors had a transcribed-to-untranscribed ratio (T/U) < 1.0, with a median ratio of 0.92 (IQR, 0.88–0.97; Wilcoxon *P* = 5.0 × 10⁻⁴). In the C>T channel, all 16 evaluable tumors had T/U < 1.0, with a median ratio of 0.93 (IQR, 0.89–0.96; *P* = 3.1 × 10⁻⁵). The observed depletion of mutations on the transcribed strand supports repair of *in vivo* lesions by transcription-coupled nucleotide excision repair (TC-NER)(^14^).

Predominant APOBEC mutagenesis was detected in a subset of HPV+SNSCC (Fig. 1a). When projected alongside TCGA HNSCC, SNSCC showed a greater diversity of APOBEC3 activity and a similar overall mutational burden (Fig. 3b, 3d). HPV+SNSCC appears to have less APOBEC3 mutagenesis on average than HPV+OPC, but 2/21 (9%) of HPV+SNSCC tumors ranked among the most enriched APOBEC-mutators in the TCGA–ICGC combined cohort (Fig. 3d). Among the three HPV-negative SNSCC tumors, two showed a dominant UV-associated mutational signature (Fig. 1a). We observed low burden of the smoking-related signature SBS4 in patients with a history of smoking(^15^), arguing against smoking as a primary etiology of SNSCC.

Ultra-deep HPV genome WGS showed that APOBEC activity extends to the viral genome (Fig. 2e, Supplementary Tables S4, S5). Highly clonal viral variants likely represent variants present in the infecting virion; however, lower-allele-fraction subclonal mutations identified in a subset of tumors likely represent somatic APOBEC-associated mutations acquired in the HPV genome in the current host (Fig. 2e, f) These lower-allele-fraction viral variants were enriched in APOBEC-compatible sequence contexts [e.g., Tp(C→G/A)pN and Tp(C→T)pH], supporting their classification as APOBEC-associated rather than inherited viral polymorphisms. Concordance of host–virus APOBEC mutagenesis has to our knowledge not previously been demonstrated as a pattern in SNSCC but has been shown in HPV+OPC(^16^).

Knowing that structural variants (SVs) are common in HNSCC(^10^), we next asked whether this extends to SNSCC (Supplementary Fig. 1a, Supplementary Tables S9–S11). We identified 221 somatic SVs, with an average of ∼10 SVs per tumor. Consistent with patterns reported in other cancers(^10^), simple deletions were the most common class of SV. We observed focal deletions affecting known tumor suppressor loci – for example, loss of *PTEN* was recurrent in multiple tumors (Supplementary Fig. 1b), suggesting that *PTEN* inactivation may play a role in some SNSCCs. Moreover, *PTEN* loss is also seen in a subset of HPV+OPC and could contribute to PI3K/AKT activation(^17^).

SVs targeting genes were common, with 41% of SVs localized to introns, exons, or other gene-regulatory elements (i.e. promoters), sometimes creating chimeric, in-frame fusion transcripts with epigenetic regulators such as *NIPBL* or *BRWD1* (Supplementary Fig. 1c, e). We also identified chromosomal catastrophe in specific tumors. In one HPV+SNSCC, copy-number oscillations and clustered breakpoints along a single chromosome arm characteristic of chromothripsis were detected, notably producing an in-frame *ERG-BRWD1* fusion product (Supplementary Fig. 1e). Analysis of whole chromosomes also identified kataegis in a separate case (Supplementary Fig. 1d, e). This suggests that as double-strand breaks were salvaged, APOBEC might have been concurrently active, peppering exposed single-stranded DNA with C→T mutations. Additionally, tumors with otherwise low mutation burden demonstrated chromoplexy-like rearrangements, i.e. showers of interrelated intrachromosomal SVs (Supplementary Fig. 1e).

## Discussion

Our findings show that HPV drives tumorigenesis in SNSCC and that non-HPV16, high-risk HPV genotypes play an underrecognized role, supporting their inclusion in routine clinical diagnostic testing. In a subset of cases, the mutagenic activities of the APOBEC3 family of anti-viral DNA cytosine deaminases accelerate tumor evolution. Unexpectedly, Sig 12 (SBS12) was the most frequently dominant mutational signature in HPV+SNSCC, and the transcriptional strand bias of its major T>C and C>T components support it as an *in vivo* mutational process. SBS12 is a mutational signature of unknown origin, primarily identified in liver, kidney, and lung cancers, with a higher prevalence observed in East Asian populations. Recent lung adenocarcinoma data suggest that SBS12 may be a relatively common mutational process in selected tumor contexts.(^18^) Detection of SBS12 in SNSCC raises the possibility of shared or convergent mutagenic mechanisms, although the underlying exposure or biological process remains unknown. To our knowledge, this is the first report documenting such a prominent presence of this signature in SNSCC. Larger whole-genome sequencing studies will be needed to define the origin and biological significance of SBS12 in SNSCC.

Viral integration events, particularly the extrachromosomal amplification of known oncogenes such as *TP63*, as well as genomic breakpoints and translocations, contribute to extensive structural variation. These structural variations collectively demonstrate the cooperative roles of both viral and non-viral axes of instability in tumor evolution. The diversity of HPV genotypes other than HPV16 provides a striking contrast to HPV+OPC, suggesting less evolutionary advantage for HPV16 outside the oropharynx.

## METHODS

### Patient Cohort and Sample Collection

Primary tumor and matched normal blood samples were collected from 21 patients diagnosed with SNSCC. Of these, 16 were from Mass General Brigham and 5 were from Vanderbilt University Medical Center. The study was conducted under IRB protocol #2022P001655. Clinicopathological features, including age, sex, smoking history, anatomical subsite, histologic subtype (keratinizing vs. non-keratinizing), and inverted papilloma association, were extracted from medical records. In addition to standard clinical assays (p16 immunohistochemistry and HPV PCR), all patients underwent gold-standard HPV genotyping performed at the U.S. National Institutes of Health (NIH) (Table 1, Supplementary Table S1). HPV detection and typing were performed using the TypeSeq v1 and NCI Carcinogenic HPV All Next Generation Sequencing (NCI CHANGeS) assays.

### Whole-genome sequencing

DNA was extracted from the tumor and blood using the DNeasy Blood & Tissue Kit (Qiagen) according to the manufacturer’s protocol. DNA quality and quantity were assessed with Qubit fluorometry and TapeStation (Agilent). Libraries for WGS were prepared using the Illumina TruSeq DNA PCR-Free Library Prep Kit (Illumina). Paired-end sequencing (151 bp) was performed on an Illumina platform to an average depth of ∼59x for tumor DNA and ∼30x for matched normal DNA. Reads were aligned to a hybrid human–HPV reference composed of human genome (GRCh37/hg19) and all HPV genomes downloaded from PaVE, using BWA-MEM (v0.7.15) with default settings.

### Ultra-deep HPV genome WGS and HPV mutation analysis

HPV DNA was captured using a custom Ion AmpliSeq panel (Thermo Fisher Scientific) with 545 primer pairs, covering overlapping amplicons across the 12 high-risk HPV types considered in this study. Whole-genome single-end sequencing was performed for all 12 HR-HPV types concurrently on the Ion PGM platform. HPV mutations were called using MuTect1 (v1.1.7) in unpaired mode using the hybrid human–HPV reference described previously(^19^). Mutations were quality-filtered and annotated by APOBEC likelihood (see Supplementary Methods).

### Mutation calling

Somatic single-nucleotide variants (SNVs) and small insertions and deletions (indels) were called using MuTect1 (v1.1.7) and Strelka2 (v2.9.10) using matched tumor/normal pairs. Mutations were quality-filtered and annotated for function and APOBEC likelihood (see Supplementary Methods). Mutations with allele fraction > 95% were considered as clonal.

### Mutational signature attribution

Non-negative matrix factorization (NMF) was applied to pooled samples from our cohort and The Cancer Genome Atlas (TCGA) to decompose mutations into respective single-base substitution signatures (see Supplementary Methods).

### Tumor ploidy

Tumor ploidy was estimated using the emcncf function from the FACETS algorithm (see Supplementary Methods).

### Mutational strand bias

We examined transcriptional strand bias in the substitution channels that comprise the SBS12 spectrum: T>C (65%) and C>T (12%). For each SNSCC tumor (n = 21), genic somatic mutations were assigned to the transcribed (T) or untranscribed (U) strand using NCBI RefSeq annotations (GRCh37/hg19), and per-patient T/U ratios were calculated for each channel. Deviation from the null expectation of T/U = 1.0 was evaluated across patients using a two-sided Wilcoxon signed-rank test.

### Identification of structural variants and copy number alterations

Somatic SVs were identified using dRanger and BreakPointer as described previously(^20^). Copy number segments were obtained via FACETS and subsequently fed into GISTIC analysis via the Broad pipeline. All HPV genomes were passed through SvABA for human-viral breakpoint detection, followed by manual review for high evidence (sufficient read depth and positional diversity) of chimeric reads suggestive of human–HPV breakpoints.

### ecDNA detection

Extrachromosomal DNA (ecDNA) amplifications were detected using Amplicon-pipeline v1.3.9. (see Supplementary Methods).

## Authors’ Contributions

**L. Zareba:** Conceptualization, analysis, methodology, investigation, visualization, writing. **S. Eldfors:** Conceptualization, analysis, methodology, investigation, visualization, writing. **M. Lin:** Analysis, methodology, visualization, writing. **M.E. Bryan:** Analysis, methodology, investigation, writing. **W.C. Faquin:** Investigation, resources, writing. **L. Mirabello:** Resources, writing. **S.K. Mishra:** Resources, writing. **J.S. Lewis Jr.:** Investigation, resources, writing. **M. Lawrence:** Conceptualization, analysis, methodology, supervision, writing. **D. Faden:** Conceptualization, resources, supervision, project administration, writing.

## Data Availability

All data produced in the present work are contained in the manuscript.

## Acknowledgments

This work was supported by NIH/NIDCR grant K23DE029811, NIH/NIDCR grant R03DE030550, and NIH/NCI grant R21CA267152 to D.L. Faden, and NIH/NCI grant R01CA262874 to M.S. Lawrence. The authors thank Harrison B. Chong and Hanjun Lee for their contributions to this study.

---

**Supplementary Figure S1. Discovery of structural variants in SNSCC.** (a) Abundance of SVs of various classes in 21 SNSCC (left). Size distributions of deletions (right). (b) GISTIC (Genomic Identification of Significant Targets in Cancer, Broad pipeline) output showing common focal copy number alterations present among 21 SNSCC, highlighting regions of focal *PTEN* copy number loss across three patients. (c) Whole-genome circos plot defining the constellation of different classes of inter-and intra-chromosomal SVs which are harbored across 21 SNSCC. Detection of an in-frame fusion between exon 24 of *NIPBL* and exon 10 of *DNAJC21* in a single patient, showcasing raw densities of both chimeric and wildtype reads at the mapped genomic breakpoints in both tumor and normal samples (top left). (d) Detection of a kataegic cluster of APOBEC-related mutations on chromosome 12 of patient 12. Radial scatter plot depicts the inter-mutational distances between mutations. (e) Tracing of structural variations to allelic and copy number imbalance in patient 8 across whole chromosomes (17, 18, 19, 20, 21). Clustered targeting of intra-chromosomal rearrangements resulting in loss of chromosome 21; focal gain on chromosome 17; oscillating copy number states on chromosome 19; as well as a chromothriptic shower of intra-chromosome 21 rearrangements, leading to an in-frame fusion between *ERG* and *BRWD1*, are highlighted.

